# Epidemic preparedness - *Leishmania tarentolae* as an easy-to-handle tool to produce antigens for viral diagnosis: application to COVID-19

**DOI:** 10.1101/2021.07.05.21260035

**Authors:** Ilaria Varotto-Boccazzi, Alessandro Manenti, Francesca Dapporto, Louise J. Gourlay, Beatrice Bisaglia, Paolo Gabrieli, Federico Forneris, Silvia Faravelli, Valentina Bollati, Gian Vincenzo Zuccotti, Emanuele Montomoli, Sara Epis, Claudio Bandi

**Author notes:** Co-first authors (Ilaria Varotto-Boccazzi, Alessandro Manenti).

## Abstract

To control future epidemics, discovery platforms are urgently needed, for the rapid development of diagnostic assays. Molecular diagnostic tests for COVID-19 emerged shortly after the isolation of SARS-CoV-2, however, serological tests based on antiviral antibody detection, revealing previous exposure to the virus, required longer developmental phases, due to the need for correctly folded and glycosylated antigens. The delay between the identification of a new virus and the development of reliable serodiagnostic tools limits our readiness for the control of a future epidemic. In this context, we propose the protozoan *Leishmania tarentolae* as an easy-to-handle micro-factory for the rapid production of viral antigens, to be used at the forefront of emerging epidemics. As a study model, we engineered *L. tarentolae* to express the SARS-CoV-2 Receptor Binding Domain (RBD) and report the ability of the purified RBD antigen to detect SARS-CoV-2 infection, with a sensitivity and reproducibility comparable to that of a reference antigen produced in human cells. This is the first application of an antigen produced in *L. tarentolae* for the serodiagnosis of a Coronaviridae infection. Based on our results, we propose *L. tarentolae* as an effective system for viral antigen production, even in countries that lack high-tech cell factories.

## Introduction

The abrupt emergence of the COVID-19 pandemic has underlined the urgent need for new discovery platforms for the rapid development of diagnostic and or monitoring tools, targeting infectious diseases [1,2]. Indeed, the effective containment of an infectious disease depends on the immediate availability and application of diagnostic tools to detect infected individuals, as soon as possible after the first confirmed cases have emerged [3]. In fact, within a few weeks after the isolation of the virus responsible for COVID-19 (now called SARS-CoV-2), the viral genome had been sequenced [4-6] and RT-qPCR diagnostic tools had been developed, for the detection of viral RNA [7]. In contrast, the development of tools for the serological diagnosis of COVID-19, used to search for antibodies against the virus in patient blood or serum, required more time, owing to the need to produce the protein antigens to be implemented in the diagnostic tests [8,9]. Protein antigens for serological diagnosis are generally produced using recombinant DNA technology, after the engineering or transfection of a cellular expression system for the production of the desired antigen [10-13]. The most widely used systems for the production of recombinant protein antigens include prokaryotes (e.g. *Escherichia coli*; [14]), yeasts (e.g. *Saccharomyces cerevisiae*; [15]), insect cells (through the baculovirus system; [16]) and mammalian cells (e.g. the human embryonic kidney cell line HEK293; [17]). All these approaches have been employed for the production of different antigens from SARS-CoV-2 [18-22] and diagnostic kits, some of which are commercially available (e.g. see [23]). However, there is a general consensus that antigens produced in mammalian cells guarantee optimal performances in the diagnosis of human viral diseases [12,24]. Indeed, the most widely used assays for the serological diagnosis of COVID-19 are based on antigens produced in human cells (e.g. the HEK293 cells). The differences in the diagnostic performances of viral antigens produced in mammalian cells, compared to those produced with other expression systems, mostly derive from the glycosylation pattern of the proteins and their folding [24]. In this context, human cells transfected with viral DNA are expected to produce protein antigens that are highly similar (or identical) to those produced by a human virus during its natural infection cycle. Diversely, prokaryotic systems, such as *E. coli*, are expected to produce non-glycosylated proteins, which would then potentially result in altered patterns in antibody detection, when used in the serological diagnosis of viral diseases. The limitations in the diagnostic performances of viral antigens produced in microbial prokaryotes are unfortunate because expression systems such as *E. coli* are easily engineered, cultured in optimized conditions in bioreactors, and very effective for high yield protein production at low cost [11,14].

An alternative microbial system for recombinant protein production is *Leishmania tarentolae*, a eukaryotic microbe that infects reptiles, but is non-pathogenic to humans and other mammals. *L. tarentolae* is classified as a BSL class I organism and has already been developed as an expression system for recombinant mammalian proteins, due to the protein glycosylation pattern guaranteed by this microbe, which mimics that of vertebrates [25-27]. Consequently, *L. tarentolae* is a very attractive system for antigen production, for both vaccine and diagnostic applications. To date, however, the use of this protozoon for the production of antigens for serodiagnosis has been very limited and has mainly been focused on antigens from protozoans of the genera *Leishmania* and *Trypanosoma*, with a sole application in the area of virology, on the hepatitis E virus [28-33].

Here, we show that *L. tarentolae* can easily be manipulated for the expression of a protein antigen from SARS-CoV-2, and that this antigen, tested against human sera, guarantees a diagnostic performance comparable to the same antigen produced in human cells. In the context of the recommendations to be prepared to face and combat emerging infections and future pandemics [3], our study provides a proof-of-principle for the potential utility of the *L. tarentolae* expression system, as an easy-to-handle tool to rapidly respond to future epidemics, to produce viral antigens and for the accelerated development of serological-based diagnostic assays and population monitoring.

## Materials and methods

### Plasmid construction and *Leishmania tarentolae* transfection

The sequence of SARS-CoV-2 Spike receptor-binding domain (RBD-SD1) was derived from the genomic sequence of the isolated virus “Severe acute respiratory syndrome coronavirus 2 Wuhan-Hi-1” released in January 2020, number MN908947 and comprises 819 nucleotides (range: 22517-23335) and 273 amino acids. The gene coding for RBD-SD1 was codon-optimized for *L. tarentolae* synthesized and subcloned into the pLEXSY-sat2 vector (Jena Bioscience) for constitutive, secreted expression. Cloning results in the incorporation of a C-terminal 6xHis-tag onto the resulting Spike fragment, which comprises the RBD and SD1 domains (hereafter Lt-RBD). The pLEXSY-sat2 vector integrates into the chromosomal 18S rRNA (ssu) locus of the *L. tarentolae* parasite. In addition, a signal sequence from *Leishmania mexicana*, that allows the secretion of the target protein into the culture medium, was added. The plasmid was cloned and propagated in *E. coli*, then the plasmid was linearized through digestion with SwaI enzyme. The host *L. tarentolae*-P10 was then transfected with the linearized plasmid by electroporation, according to the manufacturer’s procedures. Engineered strains were cultured in Brain Heart Infusion (BHI) liquid medium supplemented with porcine hemin (5µg/ml) (Jena Bioscience), penicillin, streptomycin (Pen-Strep, Jena Bioscience) and Nourseothricin (NTC, Jena Bioscience) (100 µg/ml) at 26°C in the dark under aerated conditions. For strain maintenance, cultures were diluted into fresh BHI medium twice a week.

### Evaluation of RBD-SD1 expression

Expression of the target protein was evaluated by analyzing a sample of the supernatant from ten recombinant clones of Lt-RBD by western blotting. After 72h of growth in BHI complete medium, supplemented with Nourseothricin at 26°C, *Leishmania* cultures were centrifuged 10 min at 3.000 g. Clarified supernatants were filtered using a 0.22 µm nitrocellulose membrane and concentrated at 5000 g for 30 minutes, using an Amicon ultracentrifugal filter with a MW cut-off of 10 kDa. Samples were diluted in a loading buffer 4X (Thermo Fisher), boiled for 5 min, and subsequently loaded onto a 4-20% gradient polyacrylamide gel (Bio-Rad Laboratories). Following electrophoresis, proteins were transferred onto a nitrocellulose membrane (Bio-Rad Laboratories), according to standard protocols, before blocking for 5 min at room temperature with EveryBlot Blocking Buffer (Bio-Rad Laboratories) and incubation with a 1:3000 dilution of anti-6x HisTag-HRP antibody (Thermo Fisher) in the EveryBlot Blocking Buffer for 1 h. After three washes with PBS + 0.1% *(v/v)* Tween-20 (PBS-T), the membrane was incubated for 5 min with the Clarity Western ECL Substrate (Bio-Rad Laboratories) and detected using the ChemiDoc Touch Imaging System (Bio-Rad Laboratories).

### Large-scale Expression and Purification of Lt-RBD

Recombinant strains expressing Lt-RBD were cultured for four days in complete BHI supplemented with Nourseothricin (100 μg/ml), Pen-Strep and hemin (1.25 µg/ml) at 26 °C in the dark and in constant agitation under aerated conditions. The *Leishmania* cultures (2.5 L) were centrifuged for 10 min at 3.000 g, and the supernatant was filtered on a 47 mm diameter activated carbon filter (Merck) (1 filter/0.5 L) to help remove the hemin from the growth media, prior to a second filtration step on a 0.22 µM nitrocellulose membrane (Merck). The supernatant was concentrated to approx. 100 mL using the Vivaflow 200 system (Sartorius) and compatible PES Vivaflow 200 cassette (10,000 MW cut-off), at 4°C, prior to dialysis, using a dialysis membrane with a MW cut-off of 14-16K (SpectaPor). Dialysis was carried out at 4°C in 2.6 L Binding Buffer (20 mM sodium phosphate buffer pH 7.4, containing 0.5 M NaCl and 0.01% *(v/v)* Tween-20). The dialysate was transferred into fresh buffer (lacking Tween-20) after 4 hours, and then after a third exchange, overnight at 4 °C. Lt-RBD was purified on a 5 mL HisTrap Excel column (Cytiva), pre-equilibrated with Binding Buffer. The supernatant was loaded at a flow rate of 1 mL/min using AKTA basic FPLC (Cytiva) and washed with Binding Buffer unit until the A280 nm reached baseline. Elution was carried out using a slow, stepwise (1%, 3%, 5%, 10%, 15%, 20%, 30%, 50% 70% and 100%) imidazole gradient, generated by mixing Binding Buffer with Elution Buffer (20 mM sodium phosphate buffer pH 7.4, 0.5 M NaCl and 0.5 M imidazole), eluting at each step with 4 column volumes (CVs). Following SDS-PAGE analysis, fractions containing the purified protein were pooled and concentrated to 500 μL, using an Amicon Ultra Filter unit with a MW cut-off of 10,000 (Millipore) and further purified by size exclusion chromatography on a Superdex 200 Increase 10/300 GL (Cytiva) column, pre-equilibrated in 1X PBS (Merck) at a flowrate of 0.5 mL/min at RT. Peak fractions (1 mL/fraction) were pooled, and aliquoted for conservation at -80 °C. Protein concentration was measured spectrophotometrically using a Nanodrop, using the theoretically calculated A280 nm for 1 mg/mL Lt-RBD of 1.077) and with the BCA Protein Assay Kit (Merck).

### SDS-PAGE and Western blot analyses

The expression of purified Lt-RBD was evaluated both by Coomassie staining and western blotting, as described above. In particular, the proteins were transferred to nitrocellulose membranes (Bio-Rad Laboratories) which were incubated for 5 min at room temperature with EveryBlot Blocking Buffer (Bio-Rad Laboratories) and then incubated with a 1:3000 dilution of anti-SARS/SARS-CoV-2 Coronavirus Spike Protein (subunit 1) Antibody (Thermo Fisher) in the blocking buffer for 1 h. After three washes with PBS + 0.1% *(v/v)* Tween-20 (PBS-T), the membrane was incubated with a horseradish peroxidase (HRP)-conjugated IgG anti-rabbit 1:30,000 (Thermo Fisher) in the blocking buffer for 1h. Finally, after three washes with PBS-T, the membrane was incubated for 5 min with the Clarity Western ECL Substrate (Bio-Rad Laboratories) and detected by ChemiDoc Touch Imaging System (Bio-Rad Laboratories).

### RBD production in human (HEK293-F) cells

For comparative analyses of glycosylation and molecular weight determination (see below), we recombinantly produced RBD in human cells (hu-RBD), to simulate the viral protein produced during a natural infection in humans. Briefly, hu-RBD was produced in the HEK293-F line (Invitrogen), cultivated in suspension using Freestyle medium (Invitrogen) according to [34] and then purified, following the procedure described in [35].

### Size Exclusion Chromatography - Multiangle Light Scattering (SEC-MALS) analysis

20 µl of 1 mg mL^-1^ recombinant SARS-CoV-2 RBDs hu-RBD and Lt-RBD were injected into a Protein KW-802.5 analytical size-exclusion column (Shodex) and separated with a flow rate of 1 mL min^-1^ in PBS using a high-pressure liquid chromatography (HPLC) system (Shimadzu Prominence). For molecular weight characterization, light scattering was measured with a miniDAWN multi-angle light scattering detector (Wyatt), connected to a differential refractive index detector (Shimadzu RID-20A) for quantitation of the total mass, and to a UV detector (Shimadzu SPD-20A), for evaluation of the sole protein content. Chromatograms were collected and analyzed using the glycoconjugate analysis algorithm available in the ASTRA7 software (Wyatt, using an estimated dn/dc value of 0.185 ml/g for proteins and 0.140 ml/g for glycans). Calibration of the instrument was verified by injection of 10 µl of 3 mg/l monomeric BSA (Sigma-Aldrich).

### Differential Scanning Fluorimetry (DSF)

DSF assays on recombinant SARS-CoV-2 Spike RBD samples (hu-RBD and Lt-RBD) at a concentration of 1 mg mL^-1^ in PBS buffer were performed using a Tycho NT.6 instrument (NanoTemper Technologies GmbH). Data were analyzed and plotted using the GraphPad Prism 7 (Graphpad Software).

### Serum Samples

IgG/IgM human serum sample from a convalescent COVID-19 subject was purchased from BIOIVT (cod. 368424) for use as a positive control. A depleted human serum lacking IgA/IgG/IgM (Sigma-Aldrich), a negative human serum provided by the University of Milan (UNICORN study; [23]) and a total of five pre-pandemic serum samples (year 2015), kindly provided by the University of Siena, were used as negative controls. To test the specificity of the assay a pool of heterologous sera was used (HCoV positive serum BIOIVT cod. 406910-SR1; Pertussis Antiserum 1sr WHO international standard; human Influenza antibody to A/California/7/2009 “like” (H1N1v) virus (2nd international standard; Diphteria antitoxin Human IgG -1st international standard). For comparative analyses, a cohort of one hundred human serum samples collected from asymptomatic subjects in March/April 2020, provided by the University of Milan (UNICORN study), and ten commercial human serum samples from symptomatic and asymptomatic donors (CUST-BB-19032021-1A and CUST-BB-19032021-1B-NeoBiotech), were used as high positive and low positive samples of the SARS-CoV-2 antibody response.

### In-House *Leishmania*-RBD Enzyme-Linked Immunosorbent Assay (ELISA) IgG qualification

The IgG ELISA assay was qualified for the serological detection of SARS-CoV-2 specific antibodies in human serum samples using the RBD antigen (Lt-RBD) according to International Conference on Harmonization Guidelines on Validation of Analytical Procedures (Q2 (R1)) [36].

Before starting the ELISA assay qualification experiments, a series of set-up experiments were performed to select the optimal antigen concentration. To this aim, 1, 2, 3 and 4 µg/ml coating concentrations were tested against human positive and negative sera for SARS-CoV-2 and compared with 1 μg/mL Spike-RBD (commercially available from Sino Biological, China - hereafter com-RBD) expressed and purified from HEK 293 cells. The set-up and qualification experiments were performed as described below.

ELISA plates were coated with 2 μg/mL of purified recombinant Lt-RBD. After overnight incubation at 4°C, coated plates were washed three times with 300 μL/well of ELISA washing solution containing Tris Buffered Saline (TBS)-0.05% *(v/v)* Tween 20 (TBS-T), then blocked for 1 h at 37 °C with a solution of TBS containing 5% *(w/v)* non-Fat Dry Milk (NFDM; Euroclone, Pero, Italy). Human serum samples were heat-inactivated at 56 °C for 1 h in order to reduce the risk of the presence of live virus in the sample. Subsequently, two-fold serial dilutions, starting from 1:100 in TBS-T containing 5% NFDM, were performed up to 1:51200. Plates were washed three times, as previously; then, 100 μL of each serial dilution was added to the coated plates and incubated for 1 h at 37 °C. Next, after the washing step, 100 μL/well of Goat anti-Human IgG-Fc Horse Radish Peroxidase (HRP)-conjugated antibody 1:100,000 (Bethyl Laboratories, Montgomery USA) were added. Plates were incubated at 37°C for 30 min. Following incubation, plates were washed and 100 μL/well of 3,3′, 5,5′ - Tetramethylbenzidine (TMB) substrate (Bethyl Laboratories, Montgomery, USA) was added and incubated in the dark at room temperature for 20 min. The reaction was stopped by adding 100 μL of ELISA stop solution (Bethyl Laboratories, Montgomery, USA) and read within 20 min at 450 nm. To evaluate the optical densities (OD), a SpectraMax ELISA plate (Medical Device) reader was used. A cut-off value was defined as three times the average of OD values from blank wells (background: no addition of analyte). Samples with the ODs under the cut off value at the first 1:100 dilution, were assigned as negative; samples where the ODs at 1:100 dilution were above the cutoff value, were assigned as positive.

### Qualification criteria and statistical analyses

#### 1 - Specificity

All samples were tested with a starting dilution of 1:100 in four repetitions per plate and in two different plates, to generate four reportable values per sample. The percentage of geometric standard deviation (%GSD) was calculated between the reportable values (RP) (GSD%= (eSD-1) ⨯ 100, where SD is the standard deviation between the natural logarithm (ln) results of the reportable values.

#### 2 - Precision

Precision of the assay was assessed by testing positive control as neat and pre-diluted 1:2, 1:4, 1:8, 1:16, 1:32, 1:64 and 1:128 in dilution buffer; thus, considering the sample in first well is always diluted 1:100, the effective samples starting dilution in the first well will be 1:100 1:200, 1:400, 1:800, 1:1600, 1:3200, 1:6400, 1:12800. The above-mentioned sample dilutions will be tested eight times in two different plates from one operator on day 1 (four repetitions per plate); the RP value will be calculated between two determination results from the two different plates. The same testing activity will be performed by another operator on day 2 to obtain a total of eight reportable values for each dilution (four RP values from operator 1 and four RP values from operator 2).

##### 2.1– Precision – Linearity

Linearity was evaluated by performing a linear regression analysis of the LOG2 serum dilution against the LOG2 of the Geometric Mean Titre (GMT) of all the eight reportable values of the precision experiments using the method of least squares. The coefficient of determination, y-intercept and slope of the regression line was calculated and reported.

#### 3 – Accuracy

The accuracy of the test was evaluated using the reportable values obtained for the evaluation of the Precision. According to ICH guideline Q2 [36], the accuracy can be tested using either a conventional true value or an accepted reference value. The true value was calculated from the linearity result as GMT between the 8 HP-HS RPs: the GMT result of reportable values from the neat sample were divided for the respective dilution factor to be investigated and compared with the real obtained value during testing (as GMT between the obtained RPs). Relative accuracy was evaluated by calculating the percentage of recovery on the GMT of the RP and the expected (true) titre obtained using this formula: 100*(GMT observed / GMT expected)

#### 4 – Robustness

Plates are incubated for 30 min (standard condition) with detection antibody to detect IgG specifically; in order to assess the influence of this incubation time, plates were incubated with the dilution antibody for 30 min at 37°C and two other different times: 20 min and 40 min.

The positive and negative controls were tested with a starting dilution of 1:100 in four repetitions, in two plates in two different days for each condition to obtain four reportable values.

### Comparative evaluation of SARS-CoV-2 IgG antibodies using two different RBD antigens

Specific anti-SARS-CoV-2 IgG antibodies in human sera were detected by means of an in-house RBD assay using Lt-RBD produced in *L. tarentolae* cells and com-RBD, produced from HEK 293 cells and purchased from Sino Biological. A collection of 80 human serum from asymptomatic subjects (UNICORN study; ethics committee of the University of Milan, approval number 17/20, approval date March 6, 2020) was tested at 1:100 dilution, while fifteen human sera (five high positive for SARS-CoV-2, five low positive for SARS-CoV-2 and five pre-pandemic negative for SARS-CoV-2) were tested starting from 1:100 to 1:25600 dilutions. The comparative assays were performed as described above. Briefly, ELISA plates were coated with Lt-RBD-SD1 (2 μg/ml) or commercial RBD (Sino Biological) (1μg/ml) overnight at 4°C. Then, heat-inactivated human serum samples were diluted in TBS-T, added to the wells and incubated for 1 h at 37°C. After three washes, 100 µl/well of Goat anti-Human IgG-Fc HRP-conjugated antibody diluted 1:100,000 (Bethyl Laboratories) were added and the plates were incubated at 37°C for 30 min. Plates were then washed and 100 μL/well TMB substrate (Bethyl Laboratories) were added and incubated for 20 min. Finally, 100 μL of ELISA stop solution (Bethyl Laboratories) were added to stop the reaction and the plates were read at 450 nm. A cut-off value was defined as three times the average of OD values from blank wells (background: no addition of analyte). Samples with the ODs under the cut-off value at 1:100 dilution, were assigned as negative; samples where the ODs at 1:100 dilution were above the cut-off value, were assigned as positive.

## Results

### Lt-RBD-SD1 protein sequence analysis, expression, and purification

For expression of recombinant Lt-RBD protein, residues 22517-23335 of the SARS-CoV-2 genome (GenBank:MN908947) were cloned into the pLEXSY-sat2 vector, as described in the Materials and methods section. The selected sequence encodes for the RBD domain (273 amino acids) of the SARS-CoV-2 Spike protein and also includes the SD1 subdomain of subunit S1. The inclusion of the SD1 portion was based on the evidence that this fragment is highly immunogenic [18]. Lt-RBD was successfully expressed as a secreted protein into the *L. tarentolae* culture medium, as confirmed by SDS-PAGE and Western blotting using an anti-His tag antibody (materials and methods). The protein migrated by SDS-PAGE as a single band of approximately 35 kDa, corresponding to its estimated MW; this band was visible for most of the tested clones (Figure 1(a)). The supernatant from the non-engineered *L. tarentolae*-P10 parasite was used as a negative control. The most productive clone was selected for large scale (2.5 L) expression and purification, via affinity chromatography and size exclusion chromatography (materials and methods). Large scale expression of Lt-RBD resulted in a yield of approx. 2,3 mg/L culture. The protein was identified by Western blotting using a specific anti-RBD antibody (Figure 1(b)) and judged to be pure by Coomassie blue staining (Figure 1(c)).

**Figure 1.**
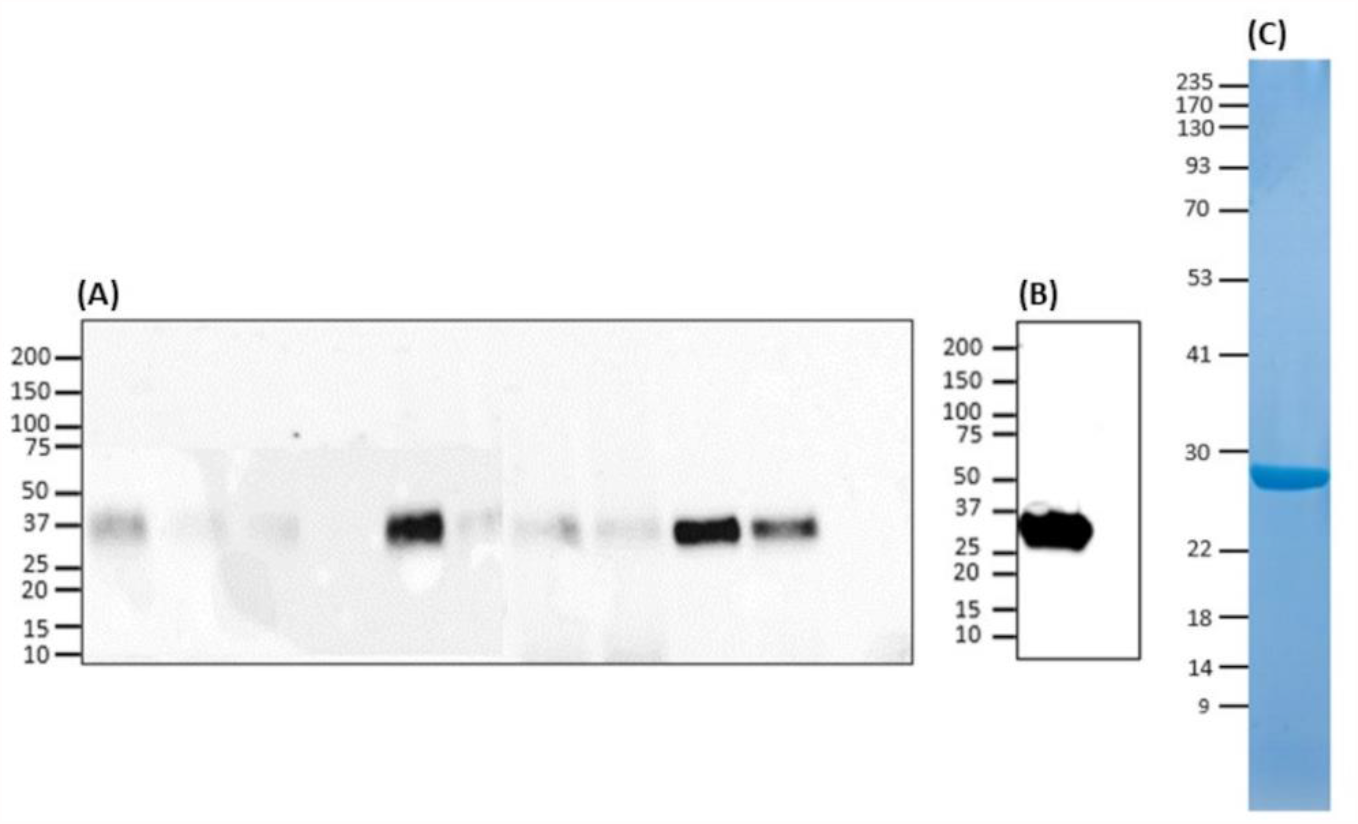
Evaluation of Lt-RBD protein production in *L. tarentolae*. a) Expression analyses of Lt-RBD in the concentrated supernatant of ten engineered *L. tarentolae* clones. A band of approximately 35 kDa is visible using an anti-His tag antibody. b-c) Analysis of Lt-RBD protein expression, purified by affinity chromatography, and confirmed by Western Blotting using an anti SARS/SARS-CoV-2 Coronavirus Spike Protein antibody (b) and by SDS-PAGE with Coomassie staining (c).

### SEC-MALS and DSF analyses

SEC-MALS analysis showed that the purified protein is monodisperse with a total molecular weight of 36.5 kDa, which is 5 kilodaltons greater than the molecular weight predicted on the basis of amino acid composition, due to the glycan moieties (Figure 2, Table 1). Molecular masses determined for hu-RBD and Lt-RBD were 31.3 and 36.5 kDa, respectively.

**Figure 2.**
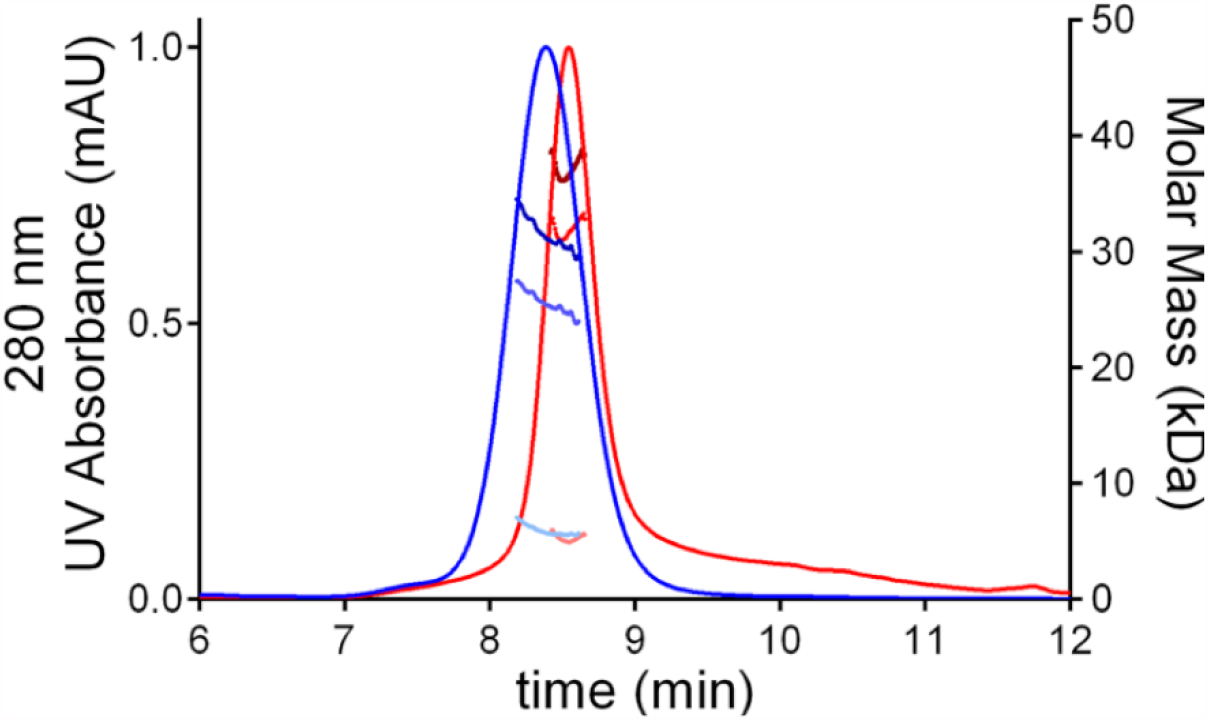
SEC-MALS chromatograms of SARS-Cov2 RBD produced in HEK293F cells, hu-RBD (blue) and in *L. tarentolae*, Lt-RBD (red). For each sample, the molar mass values associated with the glycosylated RBD molecules, the protein-only fraction, and the glycan fraction of each peak are shown using dark, intermediate, and light dots, respectively.

**Table 1.** Summary of SEC-MALS analysis

|  | hu-RBD | Lt-RBD |
| --- | --- | --- |
| *Input values for analysis* |  |  |
| dn/dc (glycoprotein) (mg/L)<sup>a</sup> | 0.185 | 0.185 |
| dn/dc (glycan) (mg/L)<sup>b</sup> | 0.140 | 0.140 |
| 280 nm Extinction coefficient (M<sup>-1</sup> cm<sup>-1</sup>)<sup>c</sup> | 1.3 | 1.39 |
| Predicted molecular weight (protein only) (g/mol)<sup>c</sup> | 25921 | 31547 |
| *Results from SEC-MALS analysis* |  |  |
| Total mass (g/mol) | 31320 ± 1180 | 36490 ± 1540 |
| Protein only (g/mol) | 25520 ± 960 | 31560 ± 1330 |
| Glycan only (g/mol) | 5794 ± 1195 | 4935 ± 1553 |

<sup>a</sup>a standard dn/dc value for proteins of 0.185 mg/L was used as suggested by the MALS manufacturer.
<sup>b</sup>a standard dn/dc value for glycans of 0.140 mg/L was used as suggested by the MALS manufacturer.
<sup>c</sup>extinction coefficients and molecular weights were estimated from protein sequences using the Expasy ProtParam tool [36,37].

The difference between the molecular masses of the two proteins is mainly due to the inclusion of the SD1 subdomain in Lt-RBD. Furthermore, their unfolding temperatures, determined by DSF analysis, are highly similar (Lt-RBD-SD1: 53.2 °C; hu-RBD: 53.4 °C), indicating that Lt-RBD is as stable as the hu-RBD protein (Figure 3).

**Figure 3.**
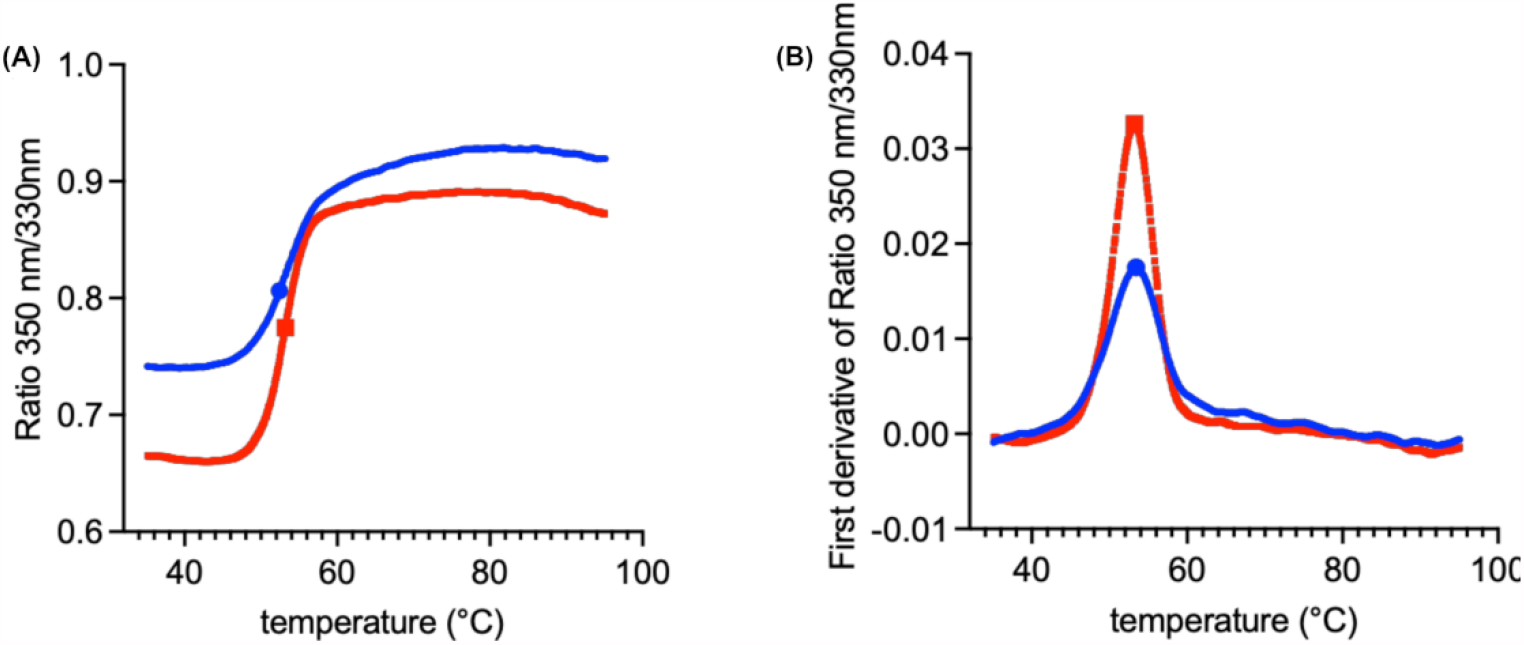
DSF traces of SARS-Cov2 RBD produced in HEK293F cells (blue) and in *L. tarentolae* (red). The raw traces (A) and the first derivative (B) of the variation of the ratio of the protein intrinsic fluorescence at 350 nm and 330 nm during a DSF experiment are shown.

### Set-up and standardization of in-house ELISA

The RBD-ELISA qualification assay was carried out, as described by [39]. Purified recombinant Lt-RBD was tested for its ability to detect specific human antibodies. The protein was evaluated using four coating concentrations (1, 2, 3 and 4 μg/mL). The optimal concentration chosen for antigen coating was 2 μg/mL and the optimal dilution for the secondary HRP conjugated anti-human IgG was 1:100,000.

#### 1 - Specificity

The specificity was determined as the ability of the assay to differentiate between similar analytes and in particular to differentiate the target analyte from non-target analytes [40]. In order to determine the specificity of the method for anti-SARS-CoV-2 IgG antibodies, positive samples for homologous and heterologous viruses/pathogens were tested. The ELISA specificity was evaluated testing the samples reported in Table 2.

**Table 2.**
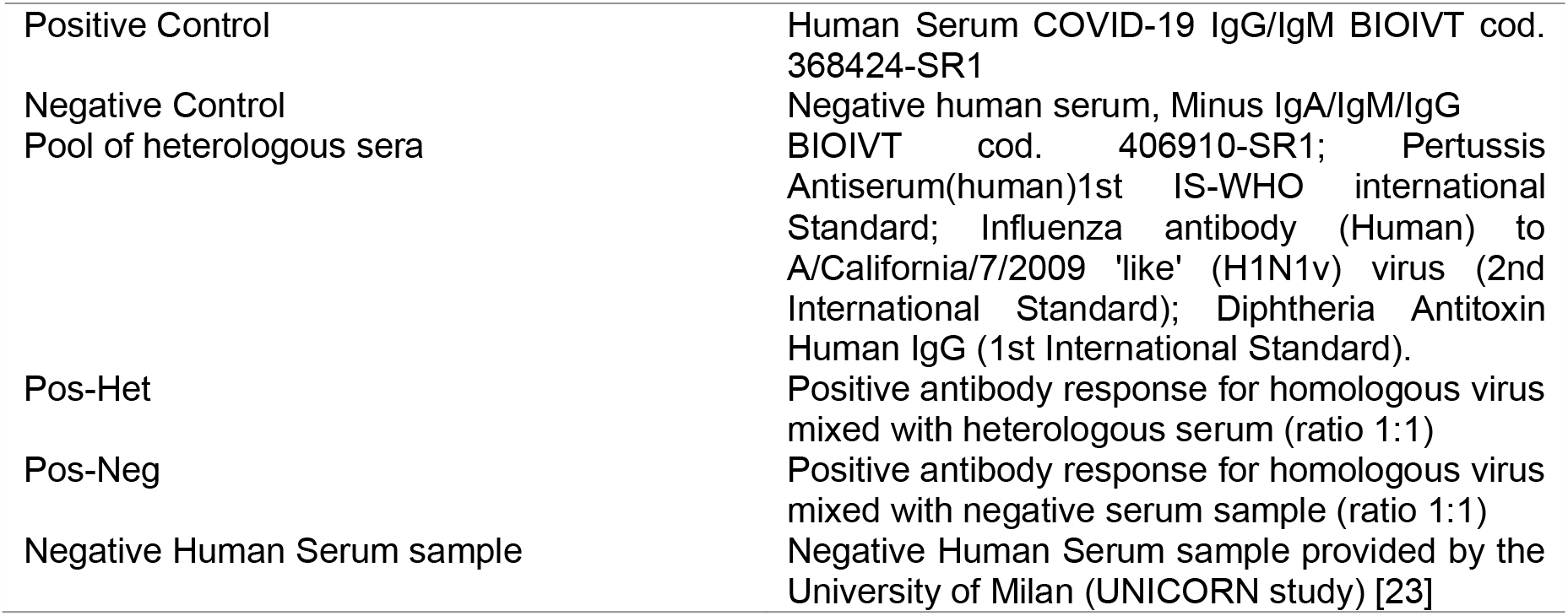
Serum samples and controls used to evaluate specificity parameters.

|  |  |
| --- | --- |
| Positive Control | Human Serum COVID-19 IgG/IgM BIOIVT cod. 368424-SR1 |
| Negative Control | Negative human serum, Minus IgA/IgM/IgG |
| Pool of heterologous sera | BIOIVT cod. 406910-SR1; Pertussis Antiserum(human)1st IS-WHO international Standard; Influenza antibody (Human) to A/California/7/2009 'like' (H1N1v) virus (2nd International Standard); Diphtheria Antitoxin Human IgG (1st International Standard). |
| Pos-Het | Positive antibody response for homologous virus mixed with heterologous serum (ratio 1:1) |
| Pos-Neg | Positive antibody response for homologous virus mixed with negative serum sample (ratio 1:1) |
| Negative Human Serum sample | Negative Human Serum sample provided by the University of Milan (UNICORN study) [23] |

The negative sample, as the heterologous sample, showed negative titres in all performed measurements of this testing series. The RP values for the sample mixtures Pos-Neg and Pos-Het samples showed a GSD% of 15.09%, which meet the acceptance criterion set at ≤50%. The positive homologous serum sample showed positive results across eight measurements in total. The negative human serum sample provided by the University of Milan (UNICORN study) showed negative values, with the starting ODs (1:100 dilution) under the cut-off value established for this validation. Overall, the results reported in Table S1 indicate that the assay is specific.

#### 2 - Precision

The precision of an analytical procedure is generally defined as the standard deviation or relative standard deviation (coefficient of variation) of a series of measurements [36,41]. The standard deviation may be evaluated at three levels: repeatability, intermediate precision and reproducibility.

##### 2.1 Precision – Repeatability

Repeatability (also called intra-assay precision) shows the precision of the assay when the test is carried out in a laboratory over a relatively short time period using the same operator and equipment. Repeatability is assessed by evaluating variation of replicates [36,40,41].

The %GSD, between four reportable values from each operator (in one experiment on the same day) from the precision experiment, was calculated and reported for each dilution, for each operator. The negative sample provided negative titre results. The positive sample showed GSD% <50% for all dilutions (Table S2). These results indicate that the assay is repeatable for intra-assay precision parameters.

##### 2.2 Precision - Intermediate precision

The purpose of intermediate precision is to determine the capacity of the assay to provide reproducible results when random events occur. Variations could include operators, equipment, different days and reagents. The intermediate precision was evaluated by performing two different assays by different operators using different sessions of analysis in the same laboratory. The %GSD between the GMT of the four reportable values from each operator from the precision experiment, was calculated and reported for each dilution. The positive sample showed GSD% <50% for all dilutions, which indicates that the assay meets the criteria for intermediate precision (Table S3).

##### 2.3 Precision – Linearity

The term “linearity” refers to the linearity of the relationship between the concentration and the assay measurement [41]. This parameter needs to be demonstrated directly for the tested analyte and to be evaluated by visual examination of a plot of signals as a function of analyte concentration [36]. The aim of linearity is to provide a model, linear or not, that is suitable to illustrate the relationship between concentration and response to the analyte [41]. The classical acceptance criteria for linearity require that the correlation coefficient of the linear regression line is close to 1, the slope showing an absolute value between 0.7 and 1.3. In the case of a significant non zero intercept, it is necessary to demonstrate that it does not have consequences on the accuracy of the assay [40]. The correlation between sample dilutions and corresponding reportable values, in the full dilutions range applied (1:100 - 1:12800), was high, in that the coefficient of determination R2 was close to 1 (0.995), while the absolute value of the slope was 1.090 (Table S4; Figure S1).

#### 3 - Accuracy

The assay quantitation range is the range within which the assay has demonstrated to have a suitable level of precision and accuracy. In detail, this range is defined by the lower and the upper sample dilutions able to provide linear and accurate results in agreement with the acceptability requirements. Although the last dilution points are almost all below the limit of detection, considering that the inter-assay and intra-assay precision are confirming the assay repeatability, the results of this validation process indicate that the assay is linear and precise along the whole range of dilutions applied in this validation resulting in a titer range between 71.0 and 14061.8, (calculated as the GMT between the neat HP-HS results in precision experiments) corresponding to the closest dilutional point of 100 and 26,500, respectively (Table S5). Although the last dilution point (1/128) is below the detection limit, the recovery value for each sample dilution was excellent.

#### 4 - Robustness

The robustness gives an indication of the assay reliability when events could occur during testing in a single laboratory. Plates are incubated for 20, 30 and 40 min in order to assess the influence of this incubation time with HRP. For the three different conditions of incubation time for antibody detection, the %GSD of the results for the positive control was <50% and the negative control showed negative results, hence the results indicate that the assay is robust (Table S6). The qualification process of this specific IgG ELISA test for SARS-CoV-2, set-up with the use of the Lt-RBD protein produced in *L. tarentolae*, fulfilled all the acceptability criteria. In particular, the assay demonstrated to be: specific, reproducible, precise, linear, accurate, and robust. Thus, the procedure is defined as reliable and valid for the serological detection of SARS-CoV-2 IgG-specific antibodies.

### Detection of IgG antibodies in human sera

The ability of Lt-RBD-SD1 to detect antibodies in comparison with the commercial RBD produced in HEK cells (com-RBD) was evaluated testing fifteen human sera, among which five high positive and five low positive for SARS-CoV-2. As negative controls, five pre-pandemic (2015) sera were included in the assay. The IgG antibody titers were calculated, and the results obtained with the two proteins were compared. As shown in Figure 4 the results, expressed as titres were highly congruent and the curves showing the trend of the results among samples exactly the same.

**Figure 4.**
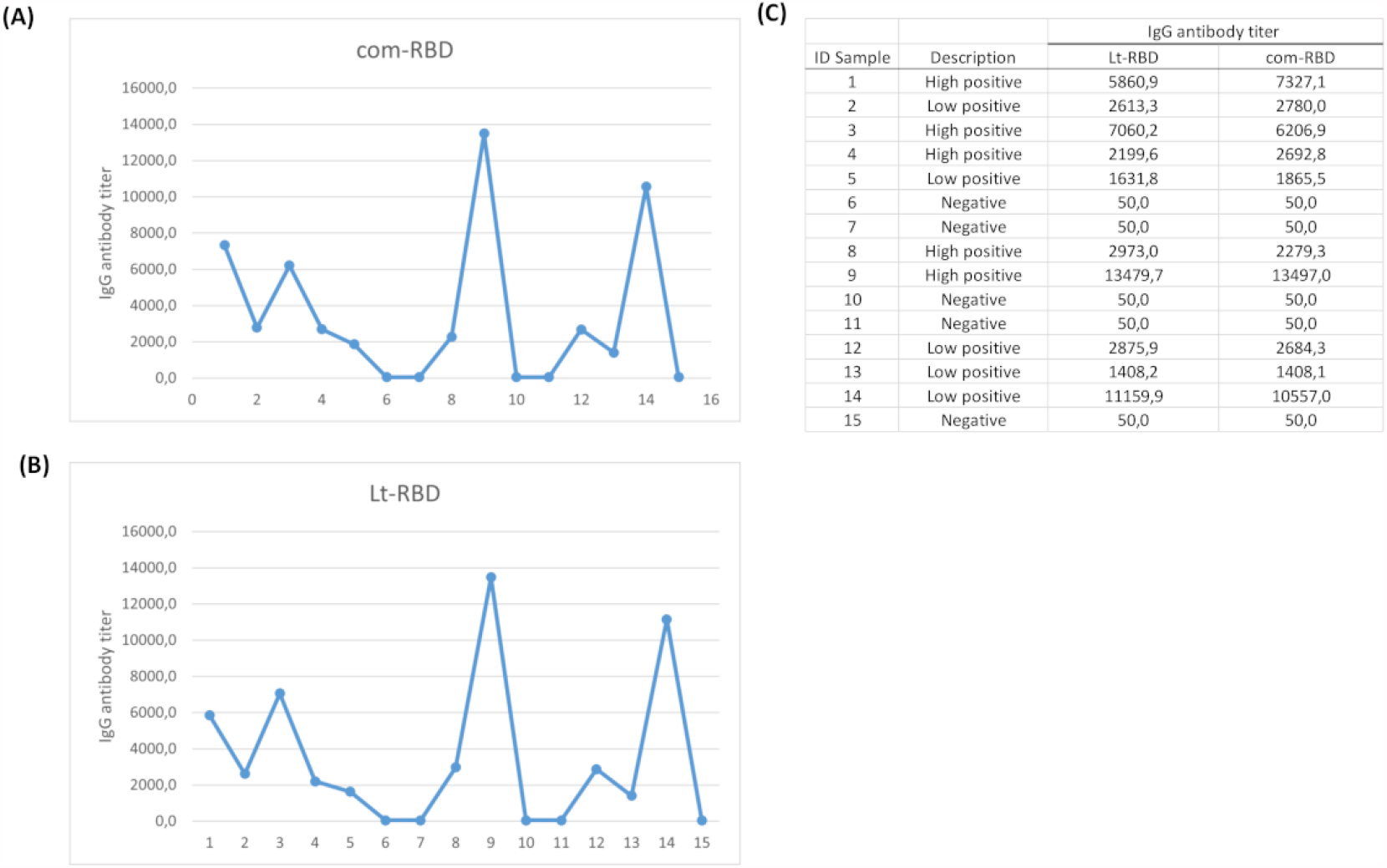
IgG antibody titers obtained in ELISA assay testing five high positive, five low positive samples for the SARS-CoV-2 antibody response and five negative sera. A,B) Curves represent the values of titers of IgG antibodies using com-RBD produced in HEK cells (A) and Lt-RBD produced in *L. tarentolae* cells (B). C) Comparison of titer values and description of the COVID-19 tested samples.

In addition, a cohort of eighty human sera, from asymptomatic COVID-19 subjects and from subjects that had no history of SARS-CoV-2 infection, tested negative by PCR in previous assays, were tested comparing Lt-RBD and com-RBD. Each serum was evaluated at 1:100 dilution by means of in-house ELISA assay. A serum sample was considered positive when its OD_450_ value was above the established cut-off (see materials and methods). The results of the qualitative analysis show an agreement in approximately 99% of the samples tested with the two different proteins (Table 3).

**Table 3:** comparative table showing the results obtained when human sera were tested using com-RBD produced in HEK cells and Lt-RBD produced in *L. tarentolae*.

| ID Number | com-RBD | Lt-RBD |
| --- | --- | --- |
| 1-2 | Positive | Positive |
| 3 | Negative | Negative |
| 4 | Positive | Positive |
| 5-6 | Negative | Negative |
| 7 | Positive | Positive |
| 8-9 | Negative | Negative |
| From 10 to 12 | Positive | Positive |
| 13-14 | Negative | Negative |
| 15-16 | Positive | Positive |
| From 17 to 22 | Negative | Negative |
| From 23 to 26 | Positive | Positive |
| 27 | Negative | Negative |
| 28 | Negative | Positive |
| 29-30 | Positive | Positive |
| From 31 to 34 | Negative | Negative |
| From 35 to 40 | Positive | Positive |
| 41 | Negative | Negative |
| 42 | Positive | Positive |
| From 43 to 51 | Negative | Negative |
| 52-53 | Positive | Positive |
| 54 | Negative | Negative |
| 55 | Positive | Positive |
| 56-57 | Negative | Negative |
| 58 | Positive | Positive |
| From 59 to 61 | Negative | Negative |
| 62 | Positive | Positive |
| From 63 to 67 | Negative | Negative |
| 68 | Positive | Positive |
| 69 | Negative | Negative |
| 70 | Positive | Positive |
| 71-73 | Negative | Negative |
| 74 | Positive | Positive |
| 75-77 | Negative | Negative |
| 78 | Positive | Positive |
| 79-80 | Negative | Negative |

## Discussion

Antigens produced in the *L. tarentolae* expression system have been assayed for the serological diagnosis of *Leishmania* infections in humans and dogs, with successful results both in terms of sensibility and sensitivity [30,31,33]. In addition, in parasites from the *Trypanosoma* genus, which are phylogenetically related to *Leishmania* (within the family Trypanosomatidae), antigens produced in *L. tarentolae* showed satisfactory performances in serological assays, for both African and American trypanosomiases [29,32]. These few studies, on the use of the *L. tarentolae* microfactory for the serological diagnosis of *Leishmania* and *Trypanosoma* infections, have been largely based on the assumption that protein production in phylogenetically related organisms would ensure proper folding and glycosylation, thus better serodiagnostic performance, in comparison with antigens produced in distantly related organisms, such as yeast. And perhaps by a positive bias towards *L. tarentolae* in people working on pathogenic species of *Leishmania* and *Trypanosoma*. Indeed, to the best of our knowledge, no studies have thus far been published on the use of antigens produced in *L. tarentolae* for serodiagnosis of infections caused by bacteria, or protozoa not belonging to the Trypanosomatidae. And there is only one application to the serodiagnosis of a viral infection, caused by the hepatitis B virus [28].

The success in the serodiagnostic use of antigens produced in *L. tarentolae* to detect *Leishmania* spp. or *Trypanosoma* spp. infections does not guarantee the efficacy of this protein microfactory as a source of antigens for infectious diseases caused by other pathogens. Using SARS-CoV-2 as a study system, our present work provides the first, sound evidence on the potential utility of *L. tarentolae* as a micro-factory to produce antigens for serodiagnosis of Coronaviridae infections. To summarize our results: i) the model antigen that we selected for this study, i.e. the RBD protein from SARS-CoV-2, was effectively produced and purified, and demonstrated to possess folding and glycosylation patterns comparable to that of the same protein produced in mammalian cells; ii) all of the tests carried out to validate an in-house ELISA assay based on Lt-RBD met the established recommendation criteria; iii) titrations on standard COVID-19 positive and negative sera showed linearity in the recorded values, in relation to the dilutions, and a concordance in the observed scores, with a perfect match in the attribution of the sera to the three reference classes (High positive; Low positive; Negative); in addition, results obtained with Lt-RBD were highly congruent with those obtained with com-RBD; iv) finally, qualitative analysis with Lt-RBD on 80 sera from a previous study led to an almost perfect match with the results obtained with com-RBD (with just one mismatch).

The gold standard for serological diagnosis of human and mammalian viruses is the use of antigens produced in mammalian cells [12,18,19,24]. However, the use of mammalian cells for the production of viral antigens presents potential limitations, in particular for rapid application and fast production in developing countries: highly specialized cell factories are indeed needed for efficient protein production in mammalian cells, and production costs are relatively high, compared to microbial-based systems. Our study underlines *L. tarentolae* as a system that is easy to manipulate and culture, providing a sound alternative for the rapid production of serodiagnostic proteins, even at the forefront of novel epidemics, e.g. in the presence of “spillover” events in tropical countries [42]. Accordingly, we emphasize that *L. tarentolae* can produce high protein yields that can easily be increased to industrial scale, by growing the parasites in bioreactors and harvesting proteins using high throughput strategies [25]. Consequently, this renders the *Leishmania* microfactory also potentially applicable in developing countries.

## Data Availability

Supplementary material is available in this Medrxiv

## Acknowledgements

The authors thank Prof. M. Bolognesi from University of Milan (Dept. Biosciences) for his suggestions and the Fondazione “Romeo ed Enrica Invernizzi”. The following reagents were produced under HHSN272201400008C and obtained through BEI Resources, NIAID, NIH: Vector pCAGGS Containing the SARS-Related Coronavirus 2, Wuhan-Hu-1 Spike Glycoprotein Receptor Binding Domain (RBD), NR-52309.

## Supplementary information

**Supplementary Table 1**: summary of specificity results for IgG ELISA; **Supplementary Table 2**: summary of repeatability: intra-assay precision results; **Supplementary Table 3**: summary of intermediate precision results for IgG ELISA; **Supplementary Table 4**: summary of linearity results for IgG ELISA; **Supplementary Table 5**: summary of accuracy results for IgG ELISA; **Supplementary Table 6**: summary of robustness results incubation time detection antibody for IgG ELISA; **Supplementary Figure 1**: correlation between Elisa titer and serum dilution.

