## Supplementary figures and images for "Epidemic preparedness - *Leishmania tarentolae* as an easy-to-handle tool to produce antigens for viral diagnosis: application to COVID-19"

### Supplementary Figure 1

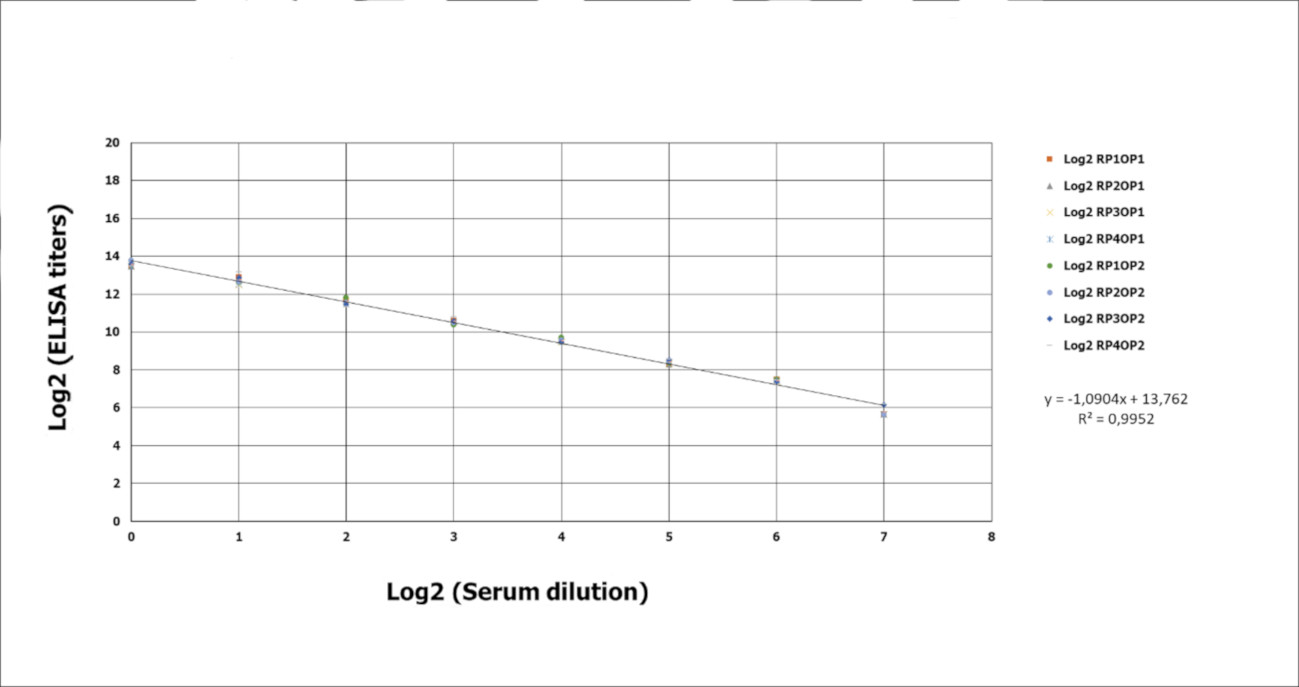
